# Treatment of malignant melanoma in certified cancer centres and its relationship to survival

**DOI:** 10.64898/2026.03.16.26347792

**Authors:** Olaf Schoffer, Daniela Piontek, Friedegund Meier, Andrew Hunter, Constanze Schneider, Andrea Sackmann, Kirsi Manz, Fabian Reinwald, Saskia Thies, Bianca Franke, Monika Klinkhammer-Schalke, Sylke Ruth Zeissig, Jochen Schmitt

## Abstract

**Background:** Hospital certification programs in Germany aim to improve the quality of cancer care. Previous research indicates that treatment in certified cancer centres leads to better overall and disease-free survival compared to non-certified hospitals for various cancers. Skin cancer, however, has not been investigated in this regard.

**Objectives:** To test the hypothesis that treatment of melanoma in a certified cancer centre is related to survival benefits.

**Methods:** Data from clinical cancer registries in Germany were analysed. The analytical sample included *n* = 47,924 patients diagnosed with malignant melanoma between 2000 and 2022. Mixed-effects Cox regression models, adjusted for demographic and clinical confounders, were used to assess overall and disease-free survival. Hazard ratios (HR) with 95% confidence intervals (CI) are reported.

**Results:** The proportion of patients treated in certified cancer centres increased over time to > 60 % from 2016 onward. Treatment in certified centres was associated with significant better overall survival (HR = 0.85, 95% CI = 0.82-0.88, p < 0.001). Results were statistically significant for stages I to III. For stage IV, the overall survival difference was not statistically significant, but subgroup analyses revealed a significant effect for cases diagnosed since 2011 (HR = 0.75, 95% CI = 0.61-0.91, p < 0.010). With regard to disease-free survival, multivariable analyses revealed better survival in certified centres (HR = 0.88, 95% CI = 0.85-0.92, p < 0.001). Similar results were observed across all subgroups stratified by stage of disease, except for stage IV.

**Conclusions:** Treatment in certified cancer centres was associated with significant survival benefits for malignant melanoma patients, suggesting that the adherence to evidence-based quality standards improves patient outcomes. The fact that one in three patients has not been treated in certified cancer centres in recent years underscores the importance of expanding access to high-quality care.

**Graphical Abstract:** 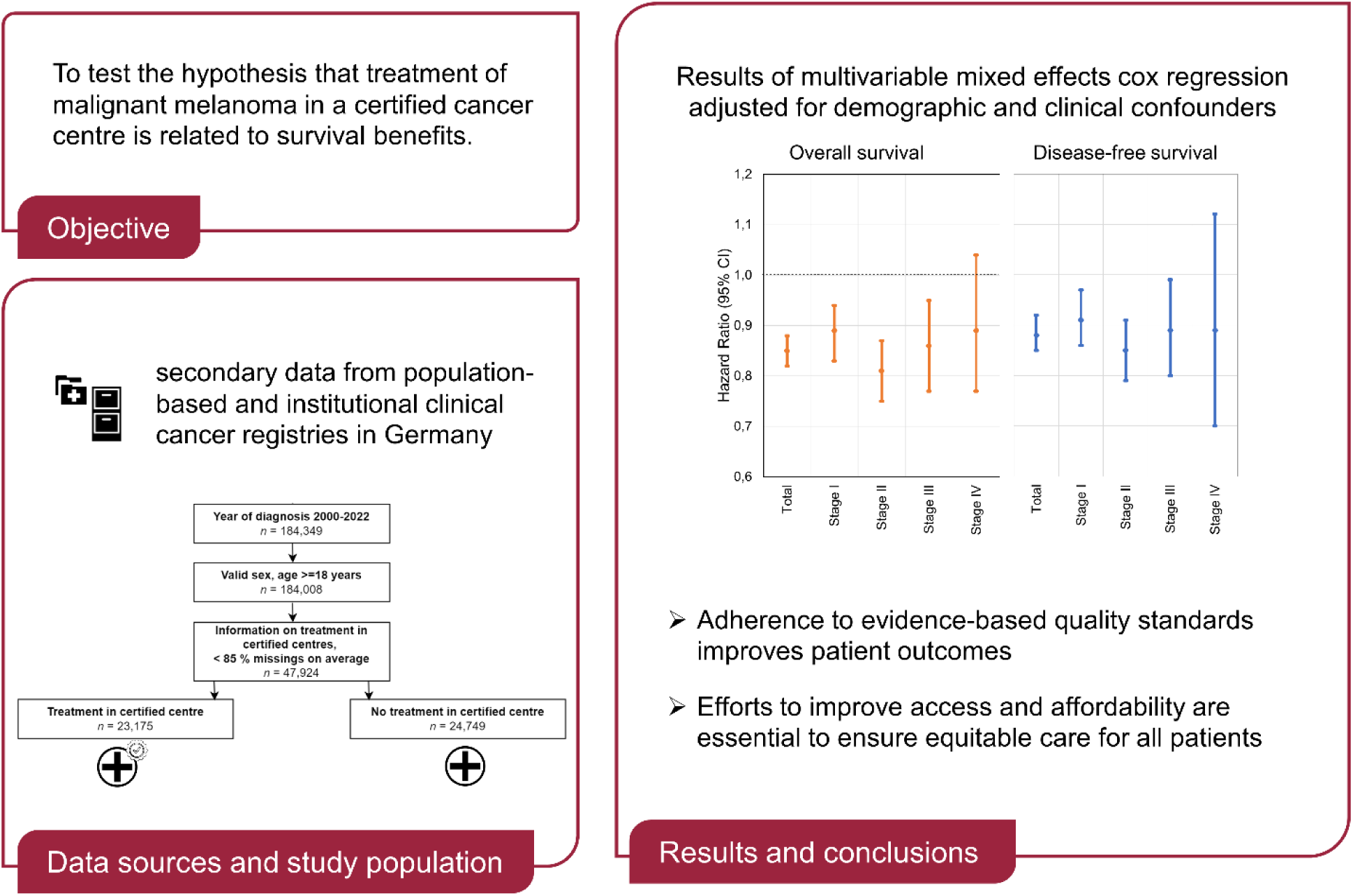

**Plain Language Summary:** *How specialized cancer centres improve survival for melanoma patients:* This study examined whether treatment in certified cancer centres in Germany improves survival outcomes for patients with malignant melanoma, a type of skin cancer. Using data from nearly 50,000 patients diagnosed between 2000 and 2022, we compared survival rates between those treated in certified cancer centres and non-certified hospitals. Certified centres adhere to strict quality standards designed to enhance cancer care. The proportion of patients treated in certified centres increased significantly over time, from 22% in 2000 to over 60% after 2016. Patients treated in certified centres had better overall survival, with a 16% lower risk of death compared to those treated in non-certified hospitals. This survival benefit was consistent across early stages of melanoma (stages I–III) and cases with unknown stages. For patients with advanced stage IV melanoma, a positive effect of treatment in certified centres was only visible for diagnoses made since 2011. Disease-free survival, which measures the time patients remain cancer-free, was also better in certified centres, showing an 12% lower risk of recurrence or progression. The findings emphasize the critical role of certified cancer centres in improving outcomes for melanoma patients. Expanding access to these centres and ensuring adherence to high-quality treatment protocols could further enhance survival rates and reduce disease recurrence, particularly for patients diagnosed at earlier stages.

**Key points:** *Why was the study undertaken?:* Existing evidence suggests that treatment in certified centres leads to better survival compared to non-certified hospitals. Using data from clinical cancer registries in Germany, this study aims to evaluate whether these benefits extend to malignant melanoma.

*What does this study add?:* Treatment in certified cancer centres is associated with significant survival benefits for malignant melanoma patients, with effects particularly pronounced for patients diagnosed at earlier stages.

*What are the implications of this study for disease understanding and/or clinical care?:* The findings underscore the importance of expanding access to certified care and ensuring adherence to high-quality treatment protocols in order to ensure equitable care for all patients.

## Introduction

Malignant melanoma is a highly aggressive form of skin cancer, accounting for a significant proportion of skin cancer-related mortality^1^. Its incidence has been steadily increasing, posing a growing public health challenge. Early detection and treatment are critical, as survival rates are closely linked to the stage at diagnosis^2^. In Germany, the survival of melanoma patients has improved over the past 20 years, especially in advanced tumour stages^3^. Despite advancements in treatment modalities, disparities in outcomes persist, emphasizing the need for optimized care delivery systems.

In Germany, hospital certification programs have been implemented to enhance the quality of cancer care. The German Cancer Society oversees one of the most comprehensive certification systems globally, encompassing over 1,000 cancer centres^4^. These programs aim to standardize care through adherence to evidence-based guidelines and multidisciplinary approaches^5^. Centres are networks of inpatient and outpatient facilities, where all disciplines involved in cancer treatment work closely together. They are required to meet specific quality indicators which are regularly monitored through a comprehensive audit process.

Certification has been associated with improved survival outcomes across various cancer types as demonstrated in large-scale studies such as the WiZen project^6–12^. In addition to its medical effectiveness, evidence suggests that centralization within the German cancer care system may also yield economic benefits^13,14^.

Existing evidence suggests that treatment in certified centres leads to better survival compared to non-certified hospitals. Using data from clinical cancer registries in Germany, this study aims to evaluate whether these benefits extend to malignant melanoma. We hypothesised that (a) melanoma patients treated in certified centres experience significantly better survival outcomes than those treated exclusively in non-certified hospitals, and (b) this survival benefit is consistent across patient subgroups.

## Patients and methods

### Data sources

Analyses are based on secondary data from population-based and institutional clinical cancer registries in Germany. The population-based registries, located in each federal state, are legally mandated to collect information on the diagnosis, treatment, and progression of cancer cases within their respective catchment areas. In addition, several comprehensive cancer centres have institutional cancer registries. Data documentation and coding follow a standardized nationwide oncological dataset. Every two years, the Association of German Tumor Centers (Arbeitsgemeinschaft Deutscher Tumorzentren e. V. (ADT) requests data from these registries to produce harmonized results on cutting edge questions in oncological health services research.

### Study population

The study cohort comprised patients with an incident diagnosis of C43 (malignant melanoma of skin) according to the International Statistical Classification Of Diseases And Related Health Problems, 10th revision, German Modification (ICD-10-GM). Moreover, the following patient level criteria had to be fulfilled: (a) malignant primary tumour, thus excluding in situ tumours, (b) histological code 8270-8790, (c) year of diagnosis between 2000 and 2022, and (d) valid sex and age >= 18 years. At registry level, information on certification had to be available with a maximum of 85% missing values on average across the observation period.

The flow chart of patient inclusion is shown in Figure 1. A total of 15 cancer registries from 11 federal states submitted data on *N* = 244,211 patients. Applying individual level inclusion criteria resulted in *n* = 184,008 patients (75.3%). Information on certification status was available from 5 cancer registries from 5 federal states, resulting in a final analytical sample of *n* = 47,924 (19.6%). Among these, *n* = 23,175 (48.4%) were treated in a certified centre, whereas *n* = 24,749 patients (51.6%) were only treated in non-certified hospitals.

**Figure 1:**
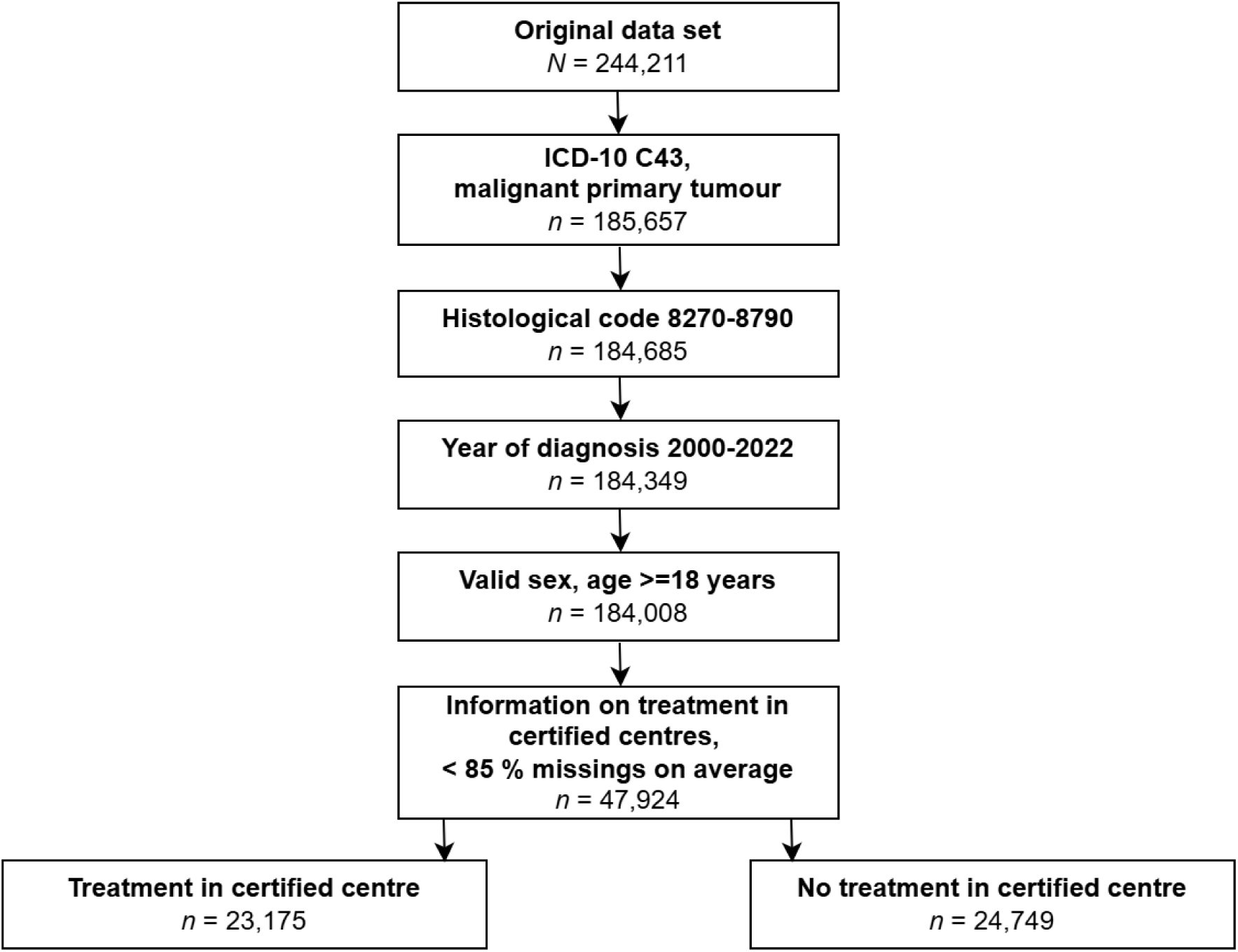
Flow chart of patient inclusion

### Treatment in certified centres

We divided our cohort into two groups: patients treated in certified centres and those treated exclusively in other hospitals. Certification status was determined using a generic, case-specific variable “centre treatment yes/no” reported by the treatment facilities. The information refers to any point in the course of treatment and indicates whether at least one part of a patient’s treatment occurred in a certified centre. However, it does not specify which part of the treatment was conducted there, whether additional treatments were performed in other hospitals or whether certification was related to the time of diagnosis or a later treatment. Even though the documentation standard refers to certification by the German Cancer Society, we could not differentiate what kind of certification the treatment facilities actually indicated. Patients with a “yes” for this variable were classified in the certified centre group, while those with a “no” or missing information were placed in the non-certified centre group.

### Confounding variables

The following variables were considered as confounders in all multivariable analyses: sex (male vs. female), age at diagnosis (18-49 vs. 50-59, 60-69, 70-79 and 80+), stage of disease according to UICC (I vs. II, III, IV and X/unknown), histological subgroup (melanoma not otherwise specified vs. lentigo maligna, superficial spreading melanoma, nodular melanoma and others) and localisation of the tumour (head and neck vs. trunk, upper extremity, lower extremity and others/missing). In addition, year of diagnosis was included as 2000-2004 vs. 2005-2009, 2010-2014, 2015-2019 and 2020-2022.

### Endpoints

The primary outcome of the study was overall survival among incident patients diagnosed with malignant melanoma. The starting point was the date of diagnosis, typically defined by the date of histological examination in cancer registry data. The observation endpoint was the date of death from any cause. The secondary outcome was disease-free survival, with date of diagnosis as starting point and observation endpoint defined as the date of local recurrence, regional lymph node recurrence, distant metastasis, unspecific recurrence, or death from any cause. Additionally, documented disease-free status during the course of the disease was required. For both outcomes, patients were right-censored if no death or relevant event was recorded until registry-specific dates of last follow-up.

### Statistical analyses

Descriptive statistics, including absolute numbers and percentages, were used to summarize categorical variables for the entire study cohort and the two study groups. Bivariate differences were assessed using Chi-squared tests. Kaplan-Meier estimation^15^ was employed to compare unadjusted survival rates between the two groups during the first five years following diagnosis. Survival probabilities (in percentages) with 95% confidence intervals (CI) were reported. Log-rank tests were conducted to determine significant differences between the study groups. To account for confounding factors and the clustered nature of the data, mixed-effects Cox regression (Cox regression with shared frailty) analyses were performed^16^. Both univariable and multivariable models were run. The latter accounted for differences in sex, age at diagnosis, histological subgroup, localisation of tumour and stage of disease (if applicable). To account for medical advancements, the grouped calendar year of diagnosis was included as confounder. A random effect for data source was incorporated to address correlated outcomes among patients documented in the same registry. Analyses were conducted for all stages combined and stratified by stage of disease. Hazard ratios (HR) with 95% CI are reported. Two-sided p-values were used, with values < 0.05 considered statistically significant. For all survival analyses, patients with a survival time of zero days were excluded (thus excluding cases registered solely based on death certificate information). Visual inspection of Kaplan-Meier plots did not suggest serious violations of the proportional hazard assumption. All analyses were performed using R version 4.4.1, employing the survminer and coxme packages for survival analyses.

### Sensitivity analyses

To assess the sensitivity of the results, stratified analyses were conducted based on sex, age group, and year of diagnosis as well as for each federal state and for the subgroup of patients with stage I to III. For stage IV, separate models were calculated for diagnoses made up to 2010 and those made since 2011 to account for the impact of newly introduced systemic therapies. Finally, analyses were restricted to population-based cancer registries, excluding data from institutional registries.

## Results

### Patient characteristics

The baseline characteristics for all patients are presented in Table 1. Compared to those not treated in certified centres, patients in the certified centre group were more likely male (*χ^2^* (1, *N* = 47,924) = 25.6, *p* < 0.001) and age 70 years or older (*χ^2^* (4, *N* = 47,924) = 124.9, *p* < 0.001). Moreover, the proportion of missing values on stage of disease was lower (*χ^2^* (4, *N* = 47,924) = 2,559.2, *p* < 0.001), the histological subgroup was more likely nodular melanoma (*χ*^2^ (4, *N* = 47,924) = 253.9, *p* < 0.001) and the tumour was more likely localised on head and neck (*χ^2^* (4, *N* = 47,924) = 66.2, *p* < 0.001). Patients from the federal state of Baden Wurttemberg were overrepresented in the certified centre group (*χ^2^* (4, *N* = 47,924) = 1,653.3, *p* < 0.001). Finally, the proportion of cases diagnosed in the earlier years of the observation period was lower, whereas it was higher in more recent years (*χ^2^* (4, *N* = 47,924) = 5,345.0, *p* < 0.001). To account for these differences, all patient characteristics were included as confounding variables in the final statistical models.

**Table 1:** Patient characteristics by treatment in certified centre.

|  | Treatment in certified centre |  |  |  |  |  |
| --- | --- | --- | --- | --- | --- | --- |
|  | yes |  | no |  | total |  |
|  | n | % | n | % | n | % |
| **Sex** |  |  |  |  |  |  |
| Male | 12,557 | 54.2 | 12,837 | 51.9 | 25,394 | 53.0 |
| Female | 10,618 | 45.8 | 11,912 | 48.1 | 22,530 | 47.0 |
| **Age group** |  |  |  |  |  |  |
| 18-49 | 4,279 | 18.5 | 5,324 | 21.5 | 9,603 | 20.0 |
| 50-59 | 3,763 | 16.2 | 3,984 | 16.1 | 7,747 | 16.2 |
| 60-69 | 5,176 | 22.3 | 5,857 | 23.7 | 11,033 | 23.0 |
| 70-79 | 6,300 | 27.2 | 6,241 | 25.2 | 12,541 | 26.2 |
| 80+ | 3,657 | 15.8 | 3,343 | 13.5 | 7,000 | 14.6 |
| **Stage of disease** |  |  |  |  |  |  |
| I | 14,292 | 61.7 | 13,966 | 56.4 | 28,258 | 59.0 |
| II | 4,757 | 20.5 | 3,752 | 15.2 | 8,509 | 17.8 |
| III | 2,207 | 9.5 | 1,295 | 5.2 | 3,502 | 7.3 |
| IV | 599 | 2.6 | 609 | 2.5 | 1,208 | 2.5 |
| X/unknown | 1,320 | 5.7 | 5,127 | 20.7 | 6,447 | 13.5 |
| **Histological subgroup** |  |  |  |  |  |  |
| Superficial spreading melanoma | 8,941 | 38.6 | 11,136 | 45.0 | 20,077 | 41.9 |
| Malignant melanoma, NOS | 5,837 | 25.2 | 5,913 | 23.9 | 11,750 | 24.5 |
| Nodular melanoma | 4,596 | 19.8 | 3,917 | 15.8 | 8,513 | 17.8 |
| Lentigo maligna | 2,530 | 10.9 | 2,650 | 10.7 | 5,180 | 10.8 |
| Others | 1,271 | 5.5 | 1,133 | 4.6 | 2,404 | 5.0 |
| **Localisation of tumour** |  |  |  |  |  |  |
| Trunk | 8,083 | 34.9 | 9,064 | 36.6 | 17,147 | 35.8 |
| Upper extremity | 5,837 | 25.2 | 6,025 | 24.3 | 11,862 | 24.8 |
| Lower extremity | 5,046 | 21.8 | 5,477 | 22.1 | 10,523 | 22.0 |
| Head and neck | 3,981 | 17.2 | 3,805 | 15.4 | 7,786 | 16.2 |
| Others/missing | 228 | 1.0 | 378 | 1.5 | 606 | 1.3 |
| **Federal state** |  |  |  |  |  |  |
| Baden-Wurttemberg | 2,924 | 12.6 | 729 | 2.9 | 3,653 | 7.6 |
| Mecklenburg Western Pomerania | 3,525 | 15.2 | 4,462 | 18.0 | 7,987 | 16.7 |
| Saxony | 9,216 | 39.8 | 11,475 | 46.4 | 20,691 | 43.2 |
| Saxony-Anhalt | 4,598 | 19.8 | 5,111 | 20.7 | 9,709 | 20.3 |
| Thuringia | 2,912 | 12.6 | 2,972 | 12.0 | 5,884 | 12.3 |
| **Year of diagnosis** |  |  |  |  |  |  |
| 2000-2004 | 1,496 | 6.5 | 5,143 | 20.8 | 6,639 | 13.9 |
| 2005-2009 | 2,656 | 11.5 | 6,486 | 26.2 | 9,142 | 19.1 |
| 2010-2014 | 5,276 | 22.8 | 5,527 | 22.3 | 10,803 | 22.5 |
| 2015-2019 | 8,057 | 34.8 | 4,537 | 18.3 | 12,594 | 26.3 |
| 2020-2022 | 5,690 | 24.6 | 3,056 | 12.3 | 8,745 | 18.2 |
| **All cases** | 23,175 | 100.0 | 24,749 | 100.0 | 47,924 | 100.0 |

NOS = not otherwise specified

### Treatment in certified centres

The proportion of patients treated in certified centres increased steadily over the observation period (Figure 2). In 2000, 21.8% of the study cohort had contact with a certified centre, with only minor changes observed until 2007. Subsequently, the proportion rose, reaching a plateau at slightly over 60% from 2016 onward. The highest proportion was recorded in 2017 at 67.9%. Over the entire observation period, less than half of all patients (48.4%) were treated in a certified centre.

**Figure 2:**
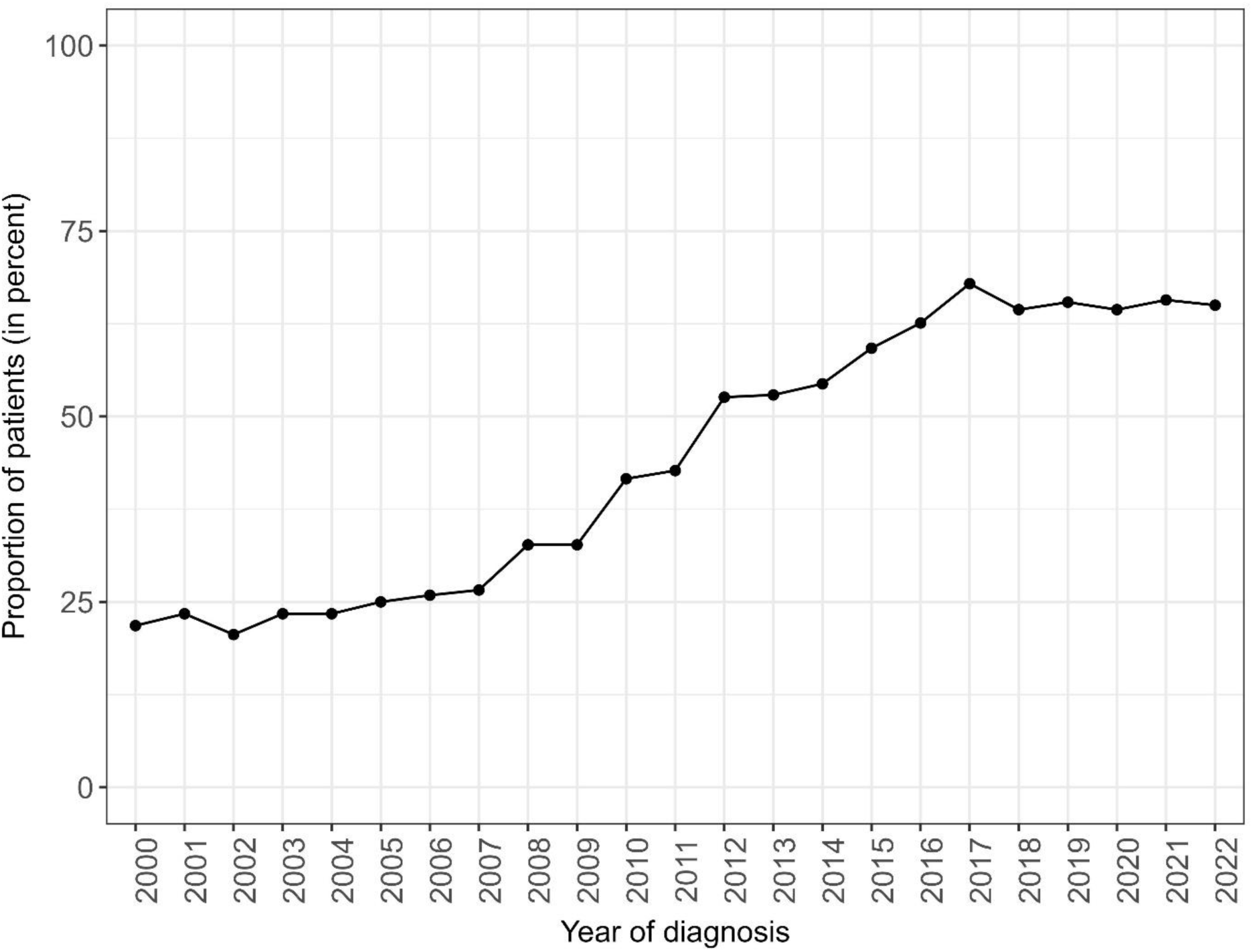
Proportion of patients treated in certified centres by year of diagnosis

### Overall survival

Based on the unadjusted Kaplan-Meier estimates, patients treated in certified centres had significantly better survival compared to those treated exclusively in non-certified hospitals (Log-rank test: (*χ*^2^ (1, *N* = 46,399) = 8.50, p < 0.010; Figure 3a). Absolute survival after 1 year was 95.7% (95% CI = 95.4%-96.0%) vs. 94.8% (95% CI = 94.5%-95.1%). After 5 years, the survival probability was 77.8% (95% CI = 77.1%-78.5%) and 75.9% (95% CI = 75.3%-76.5%) in certified centres and non-certified hospitals, respectively.

**Figure 3:**
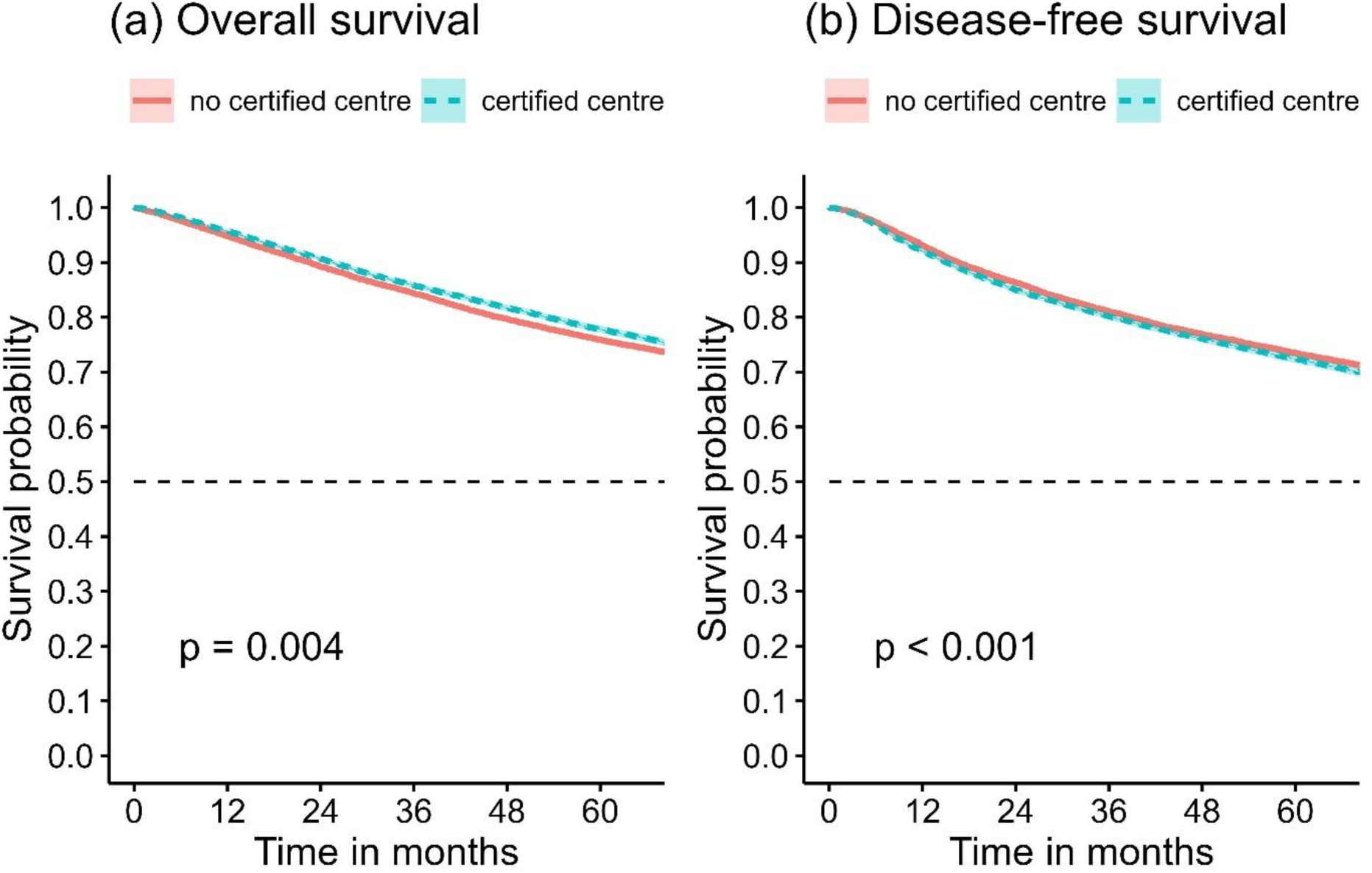
Overall (a) and disease-free survival (b) in the total study cohort by treatment in certified centres unadjusted Kaplan-Meier estimates p = significance level of log-rank test

In the univariable Cox regression, the HR for overall survival in certified centres was 0.92 (95% CI = 0.89-0.95, p < 0.001, Table 2). After adjustment for potential confounders the survival difference was even more pronounced (HR = 0.85, 95% CI = 0.82-0.88, p < 0.001). Detailed results for all covariates in the overall multivariable Cox regression model are provided in Table S1. In stratified analyses (Table 2), hazard ratios were largely consistent and statistically significant for stage I (HR = 0.89, 95% CI = 0.83-0.94, p < 0.001), stage II (HR = 0.81, 95% CI = 0.75-0.87, p < 0.001), stage III (HR = 0.86, 95% CI = 0.77-0.95, p < 0.010) and stage X/unknown (HR = 0.87, 95% CI = 0.78-0.98, p < 0.050). For stage IV, the survival difference was not statistically significant.

**Table 2:** Results of univariable and multivariable mixed effects cox regression analyses for overall survival: treatment in certified centre vs. no treatment in certified centre (reference)

| Subgroup | Univariable analysis |  |  | Multivariable analysis<sup>1</sup> |  |  |
| --- | --- | --- | --- | --- | --- | --- |
|  | p | HR | 95% CI | p | HR | 95% CI |
| Total (*n* = 46,360) | < 0.001 | 0.92 | 0.89-0.95 | < 0.001 | 0.85 | 0.82-0.88 |
| Stage I (*n* = 27,363) | 0.452 | 0.98 | 0.92-1.04 | < 0.001 | 0.89 | 0.83-0.94 |
| Stage II (*n* = 8,429) | < 0.001 | 0.89 | 0.83-0.95 | < 0.001 | 0.81 | 0.75-0.87 |
| Stage III (*n* = 3,491) | < 0.001 | 0.76 | 0.69-0.84 | < 0.010 | 0.86 | 0.77-0.95 |
| Stage IV (*n* = 1,199) | 0.094 | 0.89 | 0.78-1.02 | 0.139 | 0.89 | 0.77-1.04 |
| Stage X (*n* = 5,878) | 0.075 | 0.90 | 0.80-1.01 | < 0.050 | 0.87 | 0.78-0.98 |

Note: subgroup specific total sample sizes (*n*) after exclusion of cases with a survival time of zero days.
<sup>1</sup> adjusted for sex, age at diagnosis, histologic subgroup, localisation of tumour, (stage of disease), year of diagnosis; including random effect for data source.
HR = Hazard ratio, CI = confidence interval.

### Disease-free survival

For disease-free survival, the unadjusted Kaplan-Meier analyses showed worse survival in certified centres compared to non-certified centres for the total study cohort (Log-rank test: (*χ*^2^ (1, N = 40,972) = 12.80, p < 0.001; Figure 3b). The 5-year survival probabilities were 72.4% (95% CI = 71.7%–73.1%) and 73.5% (95% CI = 72.8%–74.2%), respectively. However, after adjusting for confounders in multivariable analyses, the survival difference reversed, indicating significant better disease-free survival in certified centres (HR = 0.88, 95% CI = 0.85–0.92, p < 0.001, Table 3). Similar results were observed across all subgroups stratified by stage of disease, except for stage IV.

**Table 3:** Results of univariable and multivariable mixed effects cox regression analyses for disease-free survival: treatment in certified centre vs. no treatment in certified centre (reference)

|  | Univariable analysis |  |  | Multivariable analysis<sup>1</sup> |  |  |
| --- | --- | --- | --- | --- | --- | --- |
|  | p | HR | 95% CI | p | HR | 95% CI |
| **Subgroup** |  |  |  |  |  |  |
| Total (*n* = 40,934) | < 0.010 | 1.05 | 1.01-1.09 | < 0.001 | 0.88 | 0.85-0.92 |
| Stage I (*n* = 25,800) | 0.291 | 1.03 | 0.97-1.09 | < 0.001 | 0.91 | 0.86-0.97 |
| Stage II (*n* = 7,888) | 0.227 | 0.96 | 0.90-1.02 | < 0.001 | 0.85 | 0.79-0.91 |
| Stage III (*n* = 3,033) | 0.083 | 0.92 | 0.84-1.01 | < 0.001 | 0.89 | 0.80-0.99 |
| Stage IV (*n* = 415) | 0.269 | 1.12 | 0.91-1.38 | 0.317 | 0.89 | 0.70-1.12 |
| Stage X (*n* = 3,798) | 0.272 | 0.92 | 0.78-1.07 | < 0.010 | 0.81 | 0.69-0.95 |

Note: subgroup specific total sample sizes (*n*) after exclusion of cases with a survival time of zero days.
<sup>1</sup> adjusted for sex, age at diagnosis, histologic subgroup, localisation of tumour, (stage of disease), year of diagnosis; including random effect for data source.
HR = Hazard ratio, CI = confidence interval.

### Sensitivity analyses

The beneficial effect of treatment in certified centres remained largely consistent in stratified analyses by sex, age, and year of diagnosis (Table S2). Over the observation period from 2000 to 2022, hazard ratios decreased, indicating increasing benefits of treatment in certified centres over time (e.g., 2000–2004: HR = 0.90, 95% CI = 0.83–0.99, p < 0.050; 2020–2022: HR = 0.64, 95% CI = 0.53–0.77, p < 0.001). Regional analyses showed comparable and statistically significant results across most federal states, except for Saxony-Anhalt. Restricting the study cohort to stages I to III also demonstrated better survival in certified centres (HR = 0.86, 95% CI = 0.82–0.89, p < 0.001). Splitting the analyses for stage IV by year of diagnosis revealed a non-significant effect for the years up to 2010, but a significant effect for the years 2011-2022 (HR = 0.75, 95% CI = 0.61-0.91, p < 0.010). A restriction of the sample to population-based cancer registries also showed a significant certification effect (HR = 0.86, 95% CI = 0.82-0.89, p < 0.001).

## Discussion

This study highlights that treatment in certified centres is associated with improved overall and disease-free survival for patients with malignant melanoma, in particular after adjusting for confounding factors. Stratification confirmed these benefits across most subgroups and suggested that the positive effects of certification may increase over time.

Our findings align with growing evidence supporting the advantages of specialized care for various cancers. For instance, the WiZen project consistently reported better survival rates in certified centres for colon and rectum^7,9^, pancreas^10^, breast^8^, lung, cervix, prostate, endometrium, ovary, head and neck, and neuro-oncological tumours^6^. While no study has compared survival rates of malignant melanoma patients treated in certified versus non-certified centres, it is argued that optimal care for complex melanoma cases is provided in specialized centres with multidisciplinary teams, tumour boards, and patient advocacy groups^17^.

Interestingly, our study found that the survival benefit was more pronounced in localized and locally advanced melanoma but was not statistically significant for stage IV disease. However, subgroup analyses revealed that the effect of being treated in a certified centre in stage IV was time-dependent, becoming evident only in diagnoses made after 2010. This finding likely reflects recent advancements in melanoma treatment. With the approval of immune checkpoint inhibitors (e.g., anti-PD-1 +/- anti-CTLA-4 therapies) and targeted therapies (e.g., BRAF/MEK inhibitors) since 2011, survival rates of stage IV patients substantially improved^18^. Certified centres are more likely to provide such cutting-edge treatments^19^ and thus exhibit better patient outcomes during periods of innovation.

The survival advantage observed in certified centres may be explained by several factors. Centres typically adopt a multidisciplinary approach, integrating the expertise of dermato-oncologists, internal oncologists, radiologists, surgeons, and even more. This ensures comprehensive decision-making, performance, and care coordination to achieve optimal patient outcomes^20^. Furthermore, centres have access to sophisticated diagnostic tools and highly skilled pathologists, enabling more precise staging and risk stratification, which supports better treatment decisions and improved prognoses^21,22^. Lastly, participation in clinical trials may further contribute to enhanced outcomes, particularly in specialized academic settings.

To ensure broader access to innovative care, greater efforts are needed to increase the proportion of patients in certified centres. In 2020-2022, melanoma patients treated in such centres had a 35% lower risk of fatal outcomes compared to those treated at non-certified hospitals. Comparable effect estimates have been shown to exhibit substantial societal relevance^12^. The fact that one in three patients has not had access to certified centres in recent years highlights significant inequalities in access to high-quality care in Germany.

A major strength of our study is the use of a large, population-based patient cohort, which allows for analyses that accurately reflect real-world cancer treatment and outcomes. The legal mandate for cancer registration in Germany ensures that the study results are highly reliable and representative^23^. Cancer registries collect data on virtually all patients within their catchment areas, minimizing the risk of selection bias. Additionally, registries track patients over time, enabling robust survival analyses and the examination of long-term effects. Compared to the WiZen project, the observation period in our study was both longer and more recent. While the data do not represent the whole of Germany, different regions in East and West Germany were included, enhancing the external validity of our results.

Our multivariable analyses included various patient- and tumour-associated factors. However, it would have been beneficial to account for additional confounders, such as socioeconomic status, patient comorbidities, and structural characteristics of hospitals. Unfortunately, due to data protection constraints, only a generic variable “centre treatment yes/no” was available. This prevented us from accessing information about the timing and type of treatment provided in certified centres, the duration of certification, or other relevant factors like hospital size, or case volume. Furthermore, the definition and completeness of the certification variable were particularly limited in the earlier years of the observation period, as treatment facilities might have documented different forms of certification or referred to time points different from the date of diagnosis. The proportion of missing values for this variable was high in the early years and decreased over time. However, this uncertainty likely leads to an underestimation of the effect of treatment in certified centres, making our analyses conservative in nature.

Although cancer registries follow standardized data documentation protocols to ensure consistency and comparability, variability in data completeness and quality persists^24^. For instance, in our sample, information on stage of disease was missing more often for patients treated exclusively in non-certified hospitals compared to those treated in certified centres (20.7% vs. 5.7%). This discrepancy likely reflects higher documentation standards in certified centres, where detailed patient information is a requirement for certification. The same consideration applies to endpoint validity. It cannot be assumed that data on vital status as well as data on recurrences and related events required, as well as follow-up data on tumour status to determine disease-free survival is complete in cancer registries. Among these, more reliable and comprehensive information is usually available from certified centres.

To eliminate potential biases and establish causal relationships between treatment in certified centres and survival, a randomized controlled trial would be ideal. However, hospital selection is often influenced by patients’ decisions and preferences, which are shaped by factors such as travel distance, recommendations, continuity of care, and insurance considerations^25^. Randomization in such a context could undermine patient autonomy and restrict access to care, making it challenging to implement this study design.

In sum, treatment in certified centres is associated with significant survival benefits for malignant melanoma patients. This finding underscores the importance of expanding access to certified care and ensuring adherence to high-quality treatment protocols. Efforts to improve access and affordability are essential to ensure equitable care for all patients. For example, outreach programs can improve cancer care by increasing awareness, promoting early detection, and enhancing access to treatment and support services, especially for underserved communities.

## Acknowledgements

The authors thank the participating clinical cancer registries for providing the data for the project: Clinical Cancer Registry Lower Saxony, Thuringian Cancer Registry, Cancer Registry Saxony, Cancer Registry of Rhineland-Palatinate in the Institute for Digital Health Data, Clinical-epidemiological Cancer Registry Brandenburg-Berlin, Bavarian Cancer Registry, Bremen Cancer Registry, Hessian Cancer Registry, Cancer Registry of Mecklenburg-Western Pomerania, Cancer Registry Saxony-Anhalt, Cancer Registry Schleswig-Holstein, Charité Berlin, CCC Tübingen-Stuttgart, OSP Stuttgart, CCC Ulm.

## Declarations

## Ethical Approval

In Germany, it is mandatory by law to prospectively report all cancer cases to the regional cancer registry. This observational study uses anonymous data from clinical cancer registries and does therefore not need ethical approval.

## Ethics Statement

Not applicable.

## Data availability statement

The data underlying this article cannot be shared publicly due to data protection regulations.

## Author contribution

Conceptualization: O.S. (Lead), D.P., F.M., J.S.; Investigation: O.S., D.P., A.H., F.M., J.S.; Data curation: D.P., A.H. (Supporting); Methodology: O.S., D.P.; Formal analysis, Software, Visualization: D.P.; Project administration: O.S., B.F.; Supervision: O.S., M.K., S.Z., F.M., J.S.; Writing - original draft: D.P. (Lead), O.S. (Supporting); Validation, Writing - review & editing: all authors; All authors read and approved the final manuscript.

