## Supplementary data for "Treatment of malignant melanoma in certified cancer centres and its relationship to survival"

Table S1: Complete results of multivariable mixed effects cox regression analyses for overall survival in the total study cohort

| | p | HR | 95% CI |
|---|---|---|---|
| **Certified centre** | | | |
| No | *Reference* | | |
| Yes | < 0.001 | 0.85 | 0.82-0.88 |
| **Sex** | | | |
| Male | *Reference* | | |
| Female | < 0.001 | 0.72 | 0.69-0.74 |
| **Age group** | | | |
| 18-49 | *Reference* | | |
| 50-59 | < 0.001 | 1.49 | 1.36-1.62 |
| 60-69 | < 0.001 | 2.39 | 2.22-2.58 |
| 70-79 | < 0.001 | 4.83 | 4.49-5.19 |
| 80+ | < 0.001 | 11.45 | 10.61-12.35 |
| **Stage of disease** | | | |
| I | *Reference* | | |
| II | < 0.001 | 1.84 | 1.75-1.93 |
| III | < 0.001 | 2.90 | 2.73-3.08 |
| IV | < 0.001 | 7.50 | 6.95-8.09 |
| X/unknown | < 0.001 | 1.54 | 1.46-1.63 |
| **Histological subgroup** | | | |
| Malignant melanoma, NOS | *Reference* | | |
| Lentigo maligna | < 0.001 | 1.25 | 1.17-1.34 |
| Superficial spreading melanoma | < 0.001 | 1.41 | 1.32-1.50 |
| Nodular melanoma | < 0.001 | 1.33 | 1.22-1.45 |
| Others | 0.933 | 1.00 | 0.94-1.06 |
| **Localisation** | | | |
| Head and neck | *Reference* | | |
| Trunk | < 0.001 | 0.87 | 0.82-0.92 |
| Upper extremity | < 0.001 | 1.37 | 1.21-1.55 |
| Lower extremity | < 0.001 | 0.88 | 0.84-0.93 |
| Others/missing | < 0.001 | 0.82 | 0.77-0.86 |
| **Year of diagnosis** | | | |
| 2000-2004 | *Reference* | | |
| 2005-2009 | 0.939 | 1.00 | 0.95-1.05 |
| 2010-2014 | 0.590 | 1.01 | 0.96-1.07 |
| 2015-2019 | 0.959 | 1.00 | 0.94-1.06 |
| 2020-2022 | 0.951 | 1.00 | 0.90-1.10 |

HR = Hazard ratio, CI = confidence interval, NOS = not otherwise specified.

Table S2: Results of multivariable mixed effects cox regression analyses for overall survival for different study subgroups (Hazard ratios and 95% confidence intervals for certified centres compared to non-certified centres)

| Subgroup | p | HR | 95% CI |
|---|---|---|---|
| **Sex** | | | |
| Males | < 0.001 | 0.87 | 0.83-0.92 |
| Females | < 0.001 | 0.81 | 0.76-0.86 |
| **Age group** | | | |
| 18-49 | < 0.010 | 0.82 | 0.71-0.95 |
| 50-59 | < 0.001 | 0.77 | 0.68-0.88 |
| 60-69 | < 0.001 | 0.84 | 0.77-0.91 |
| 70-79 | < 0.001 | 0.82 | 0.77-0.88 |
| 80+ | < 0.010 | 0.90 | 0.84-0.97 |
| **Year of diagnosis** | | | |
| 2000-2004 | < 0.050 | 0.90 | 0.83-0.99 |
| 2005-2009 | 0.051 | 0.93 | 0.86-1.00 |
| 2010-2014 | < 0.001 | 0.85 | 0.79-0.92 |
| 2015-2019 | < 0.001 | 0.76 | 0.70-0.83 |
| 2020-2022 | < 0.001 | 0.64 | 0.53-0.77 |
| **Federal state** | | | |
| Baden-Wurttemberg | < 0.010 | 0.79 | 0.69-0.92 |
| Mecklenburg Western Pomerania | < 0.001 | 0.76 | 0.69-0.84 |
| Saxony | < 0.001 | 0.84 | 0.79-0.88 |
| Saxony-Anhalt | 0.179 | 0.94 | 0.86-1.03 |
| Thuringia | < 0.001 | 0.80 | 0.71-0.90 |
| **Stage of disease** | | | |
| I-III | < 0.001 | 0.86 | 0.82-0.89 |
| **Stage IV** | | | |
| 2000-2010 | 0.296 | 1.14 | 0.89-1.45 |
| 2011-2022 | < 0.010 | 0.75 | 0.61-0.91 |
| **Type of registry** | | | |
| Population-based only | < 0.001 | 0.86 | 0.82-0.89 |

Note: The full set of covariates is included in the models but not shown in the table.

HR = Hazard ratio, CI = 95 % confidence interval.
